## Supplementary Materials for "Sub-national modelling of surveillance sensitivity to inform declaration of disease elimination"

#### A Scenario tree probability definitions

A “best” estimate and upper/lower limits are specified for each stepwise probability within the scenario tree defined in *Figure 1*. These values are then used to define a Beta distribution (between 0 and 1) in which 95% of the density falls within the given bounds. Subsequent calculations are then based on 1,000 draws from each distribution, to propagate this uncertainty into the final sensitivity estimates.

##### Acute Flaccid Paralysis (AFP) Surveillance

Given the presence of an infected individual in a LGA, the probability that a positive outcome will be observed through *AFP surveillance* is calculated as the combination of:

1. Probability that infection is an AFP case
2. Probability of notification
3. Probability of adequate stool sample
4. Probability of positive result given infection (sensitivity of test)

###### Symptomatic infection

The probability that an infection develops into clinical AFP symptoms is assumed based on an average rate of 1 clinical case per 190 WPV infections, with a 95% uncertainty interval from 1 per 150 to 1 per 250 (2,3).

###### Case notification

AFP in children is a visible, debilitating condition that causes significant concern and motivates prompt care-seeking, as illustrated by a study from Ethiopia finding that 69% of parents sought care from a public health facility for their child with AFP (4). In Nigeria during this period of time, there was intensive messaging around recognising and reporting AFP due to the WPV elimination efforts and target notification rates for surveillance (5). We therefore work from the assumption that, where absolute AFP notification rates are high (i.e. meeting the WHO threshold), the probability of notification to a public health facility is high and at least more likely than not. The value is, however, uncertain due to unobserved barriers that exist between seeking and actually *accessing* the appropriate care. We therefore set a beta distribution for this parameter which predominantly extends between 0.6 and 0.99, with highest density at 0.9.

It is not clear at what point a further increase in the rate of AFP cases reported translates to an increased probability that an incident case would be notified. For this analysis, we can only interpret this relative to the defined target level of 2 AFP cases reported for every 100,000 children under 15. Sampled values from the above distribution are therefore scaled with respect to this target, with values less than 2/100,000 scaled between 0 and 1 and values greater than 2/100,000 assigned a value of 1.

##### Stool sample collection

The probability of an adequate stool sample being collected from the notified case (defined as two samples collected at least 24 hours apart, stored to an appropriate temperature and received at the lab within 72 hours (6)) is estimated based on the previous 12 months of observations, per LGA. The proportion of prior AFP observations which are recorded as having had adequate stool collected is calculated per LGA, along with a corresponding binomial 95% confidence interval.

##### Lab test sensitivity

The probability that a test of the infected sample will yield a positive result is simply the standard diagnostic sensitivity of the test. This is assumed to be 97% with an uncertainty interval from 95% to 99.9%, informed by the cited study from Gerloff et al. (2018).

#### Environmental (ENV) surveillance

Given the presence of an infected individual in a LGA, the probability that a positive outcome will be observed through *ENV surveillance* is calculated as the combination of:

1. Probability that the infected individual resides within the catchment of a sampling site
2. Probability that a sample is collected while the individual is shedding virus
3. Probability that the sample collected is of adequate quality for virus detection
4. Probability that the sample tests positive given that virus is present (sensitivity of test)

##### Catchment area

The probability that the infected individual resides within the catchment of environmental surveillance is defined according to the positioning of active sampling sites within each LGA relative to the population. An “active” sampling site is defined as a site which has reported any sample collection during the month in question. We assumed a 5km catchment radius for each site, but explored assumptions of 2km and 10km in sensitivity analyses.

The defined catchment areas around each site were then aggregated per LGA to align with the modelled units (ie. LGA). The proportion of the total LGA population that falls within the catchment of any site in that LGA is calculated by overlaying these boundaries onto a Worldpop population count raster (100m resolution) and summing the pixel-level counts covered. Upper and lower uncertainty limits are defined as +/-15% of this proportion.

##### Sampling frequency

The probability that the infected individual is shedding at the time of sampling is estimated based on a 30 day month and assuming that an infected individual sheds virus for a Poisson-distributed number of days after onset of paralysis, with a mean of 16 (7,8). It is assumed that there is an equal chance of sampling on every day of the month, which depends on the sampling frequency. Incorporating the poisson-distributed uncertainty of shedding period and uniform uncertainty as to which days of the month are sampled, the probability that sampling coincides with shedding for one individual was estimated by simulation for a frequency of one (minimum) to thirty (maximum) sampling days per month.

This process was iterated 1,000 times, drawing 100 simulated individuals each time, to construct a distribution for the probability that sample collection on a random day would coincide with the period of shedding, given a certain frequency of sampling per month. The mean and 95% quantile intervals across repeated simulations are used to define a distribution from which to draw during sensitivity estimation.

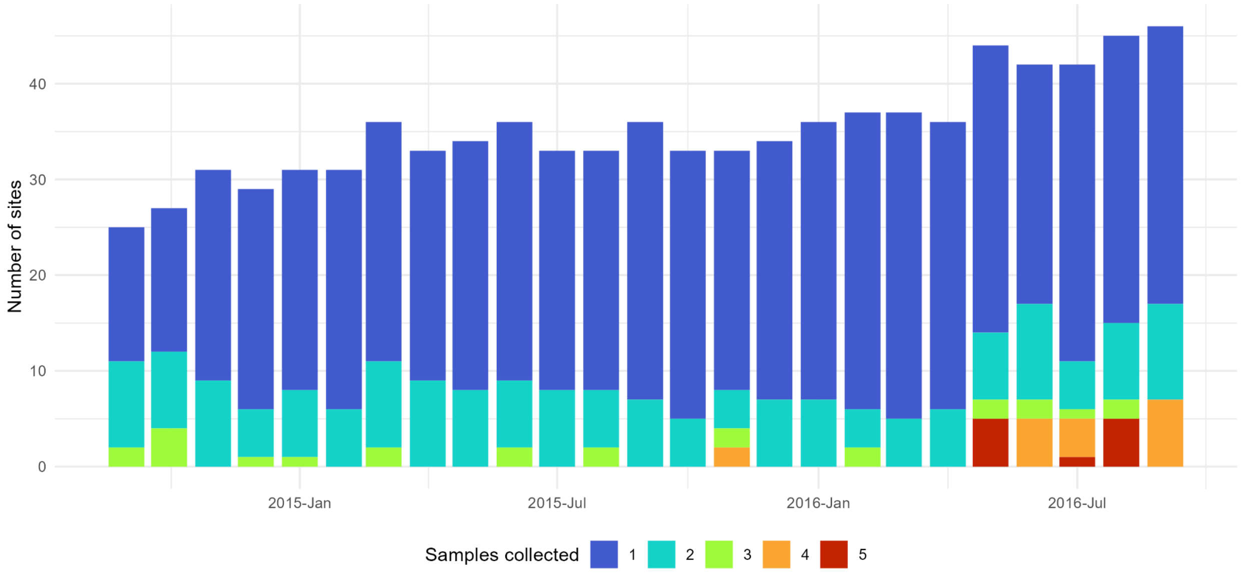


***Figure S1:*** *Frequency of sample collection per ES site per month, from Sep 2014 - Sep 2016. From May 2016 frequency increased at many sites, alongside an increase in the number of active sites overall.*

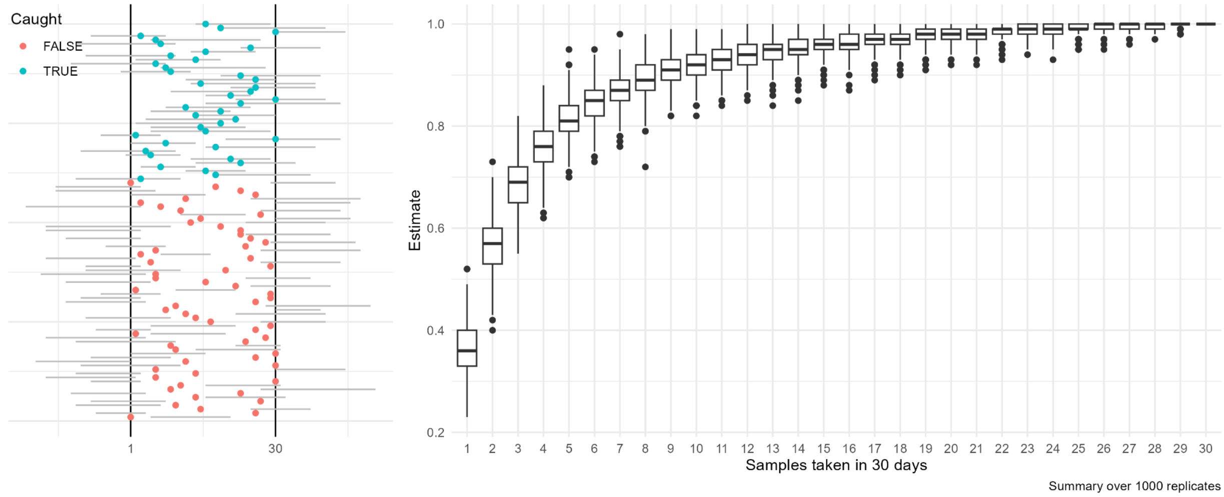


***Figure S2: (A)** Example simulations of shedding durations for a single infected individual within an ES catchment in a given month (30 days), and the coincidence of a random monthly sample collection. **(B)** Simulated probabilities of sample collection coinciding with the period of shedding of a single infected individual within the catchment, for assumed sampling frequency from 1-30 per 30 days. These distributions are then used to determine the scenario tree probability of sample collection during shedding, given the recent frequency of sampling observed at sites in a certain LGA.*

##### Sampling quality (NPEV detection)

We assume that the probability that a collected sample is of adequate quality to detect virus is indicated by the detection rate of *non-Polio enterovirus* (NPEV) among previous samples within the same LGA. A rate of at least 50% NPEV detection is considered to indicate a functioning surveillance system (9). The overall proportion of collected samples in which NPEV was detected over the previous 12 months in a given LGA (across any number of sites) is calculated along with a 95% binomial confidence interval. Sampled values from this distribution are scaled with respect to the threshold rate of 50%, i.e. values less than 50% are scaled between 0 and 1 while values greater than 50% are assigned a value of 1. In doing this, we are assuming that sensitivity is *maximised* with respect to sampling quality when this average rate at least meets the WHO threshold. The sensitivity in LGAs with lower rates of NPEV detection is then assumed to scale proportionally with how far below the threshold they fall.

##### Lab test sensitivity

The probability that the collected sample will yield a positive result given the presence of shed poliovirus is assumed to be 0.9, with uncertainty interval from 0.7 to 0.99. This is informed by the same study cited for stool sample analysis (Gerloff et al. 2018), as the assay is the same. We incorporate some additional uncertainty to account for the unknown impact of degradation, but expect this to be small as poliovirus is persistent in the environment/fresh water (hence the use of environmental surveillance). The value is currently fixed between LGAs over time, however in reality it could be influenced by the quality of the water sample and the conditions in which it was collected.

### B Surveillance activity 2016-2020

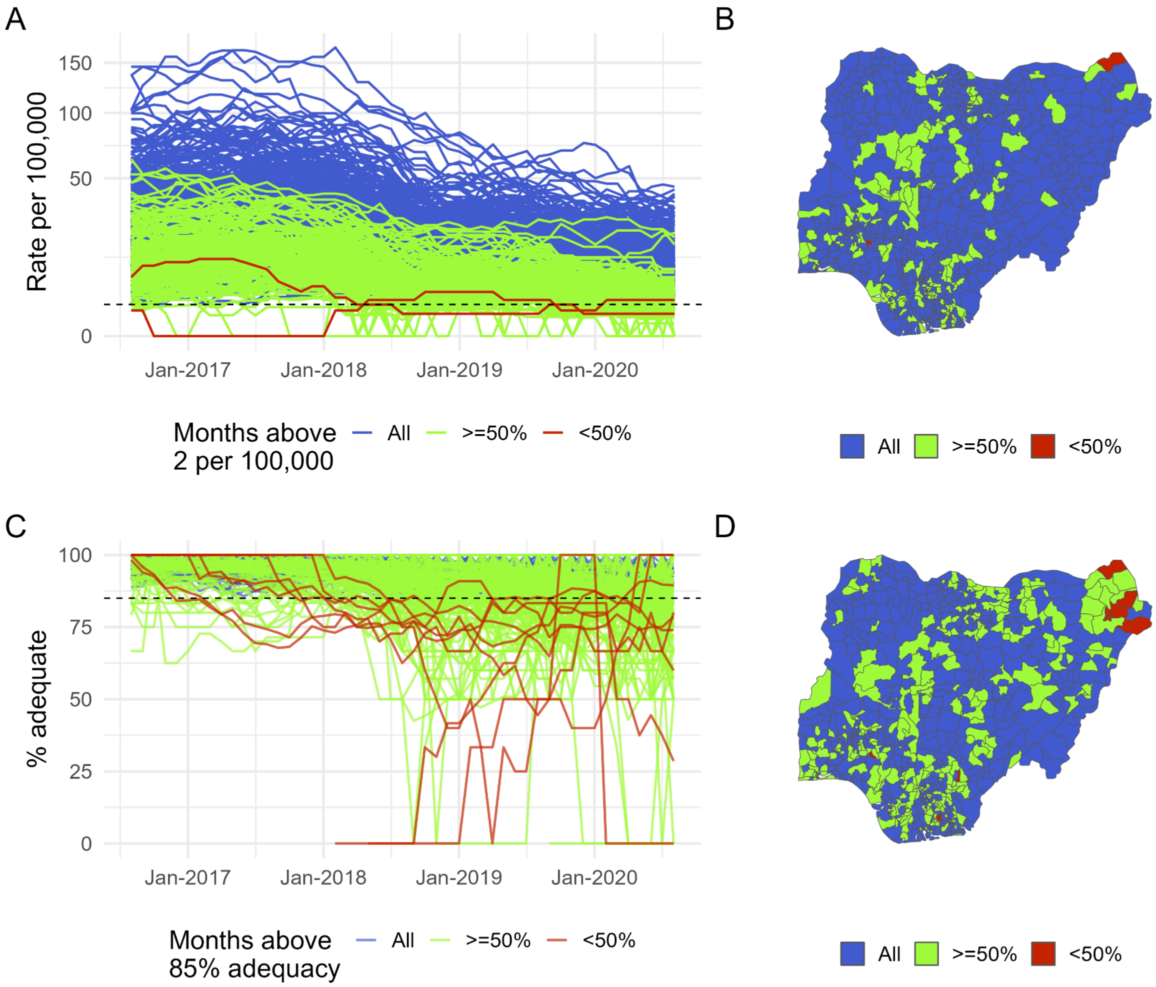


***Figure S3:*** *Equivalent to Figure 3 in the main text, but for the period 2016-2020.*

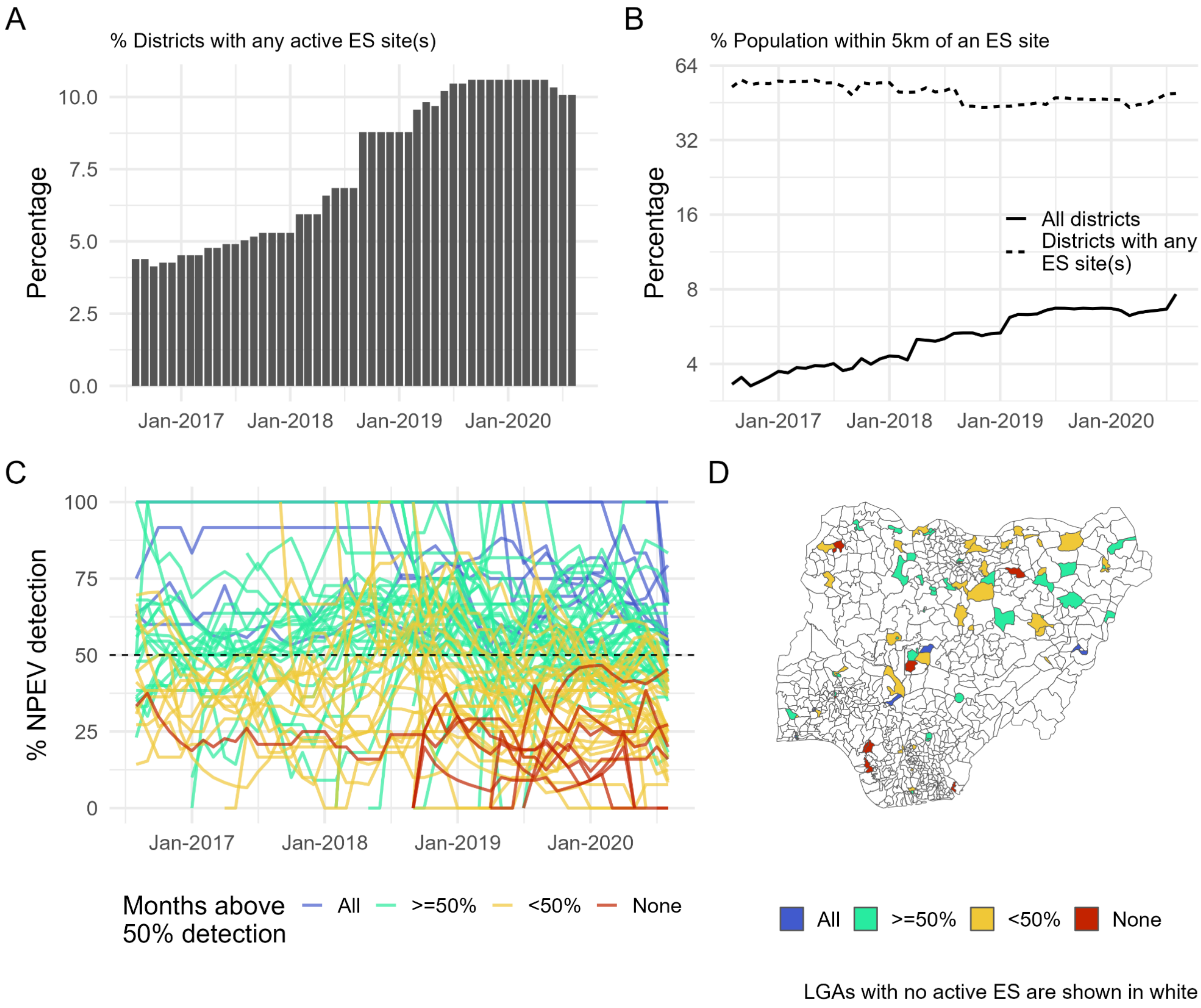


***Figure S4:*** *Equivalent to Figure 4 in the main text, but for the period 2016-2020.*

### C Sensitivity analyses

#### ES catchment radius

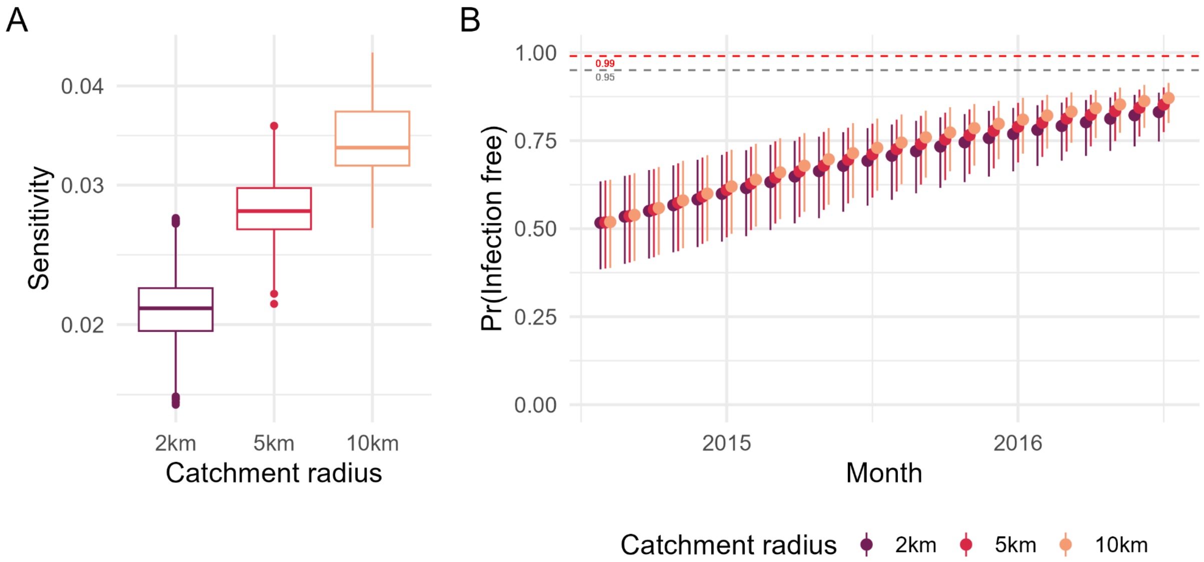


***Figure S5 (A) (B):*** *Resulting probabilities of freedom from infection given different assumptions for the catchment radius around ES sites. Expanding the radius up to 10km (highly unrealistic) yields marginal gains compared to the primary assumption of 5km.*

#### Design prevalence

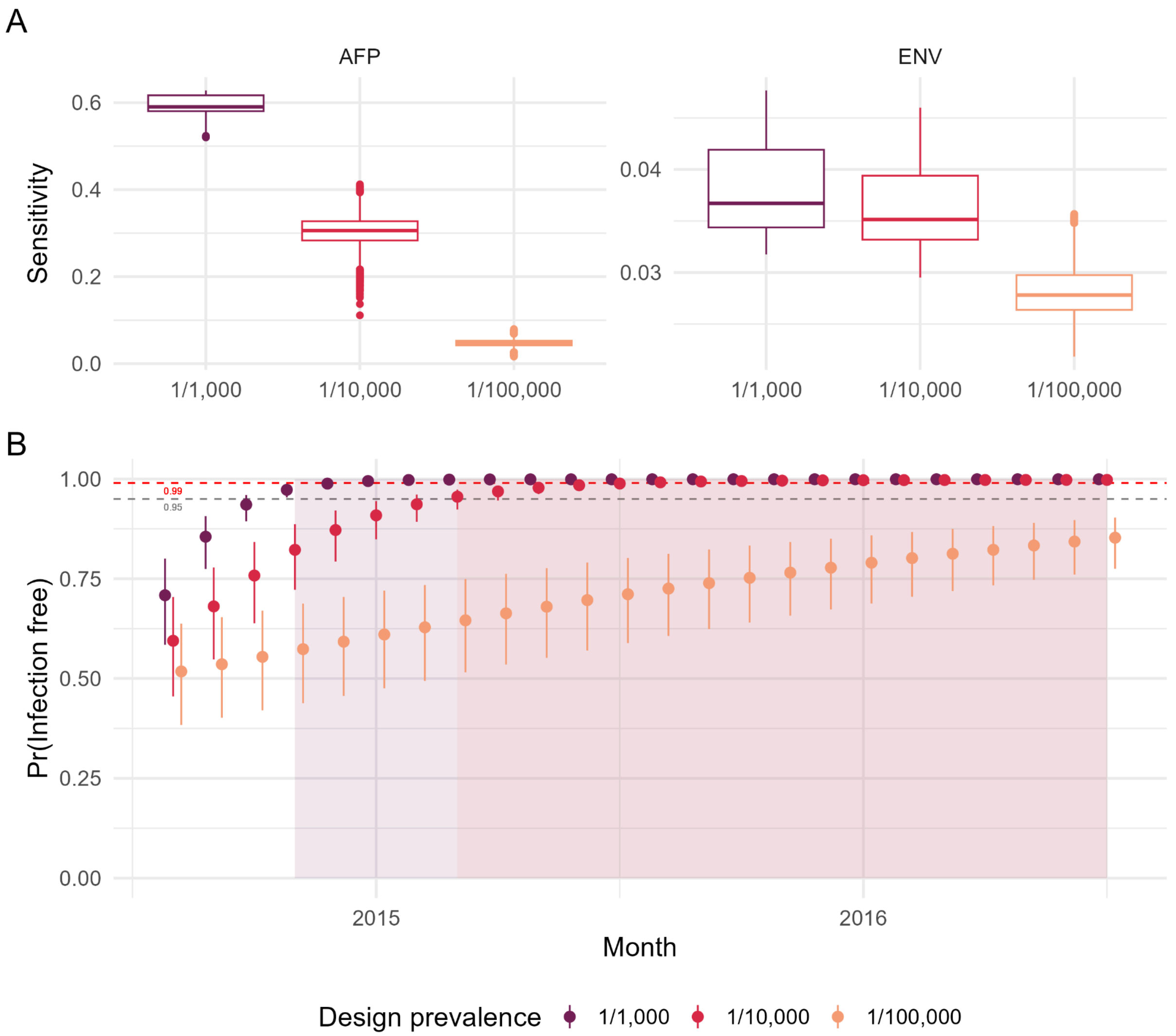


***Figure S6 (A):*** *Estimated sensitivity of the two surveillance components across all LGAs, varying the assumed design prevalence (the minimum WPV1-infection prevalence to be detected) from 1 per 1,000 to 1 per 100,000. In particular for AFP surveillance, sensitivity is substantially higher if we only require the system to detect as low as one infection per 1,000 population. The difference is smaller for ENV surveillance, as it is able to detect infection directly as opposed to only clinical disease.* ***(B):*** *Resulting probabilities of freedom from infection given different assumptions for the design prevalence. Shaded areas highlight where the probability of freedom from infection has exceeded 0.95. Assuming a higher (i.e. less stringent) design prevalence results in a much quicker accumulation of evidence that infection is below this specified level, reaching the elimination threshold within only four months for 1 per 1,000 and around 12 months for 1 per 10,000. This is not consistent with what we know in retrospect, that transmission persisted undetected during this time.*

#### Prior probability of FFI

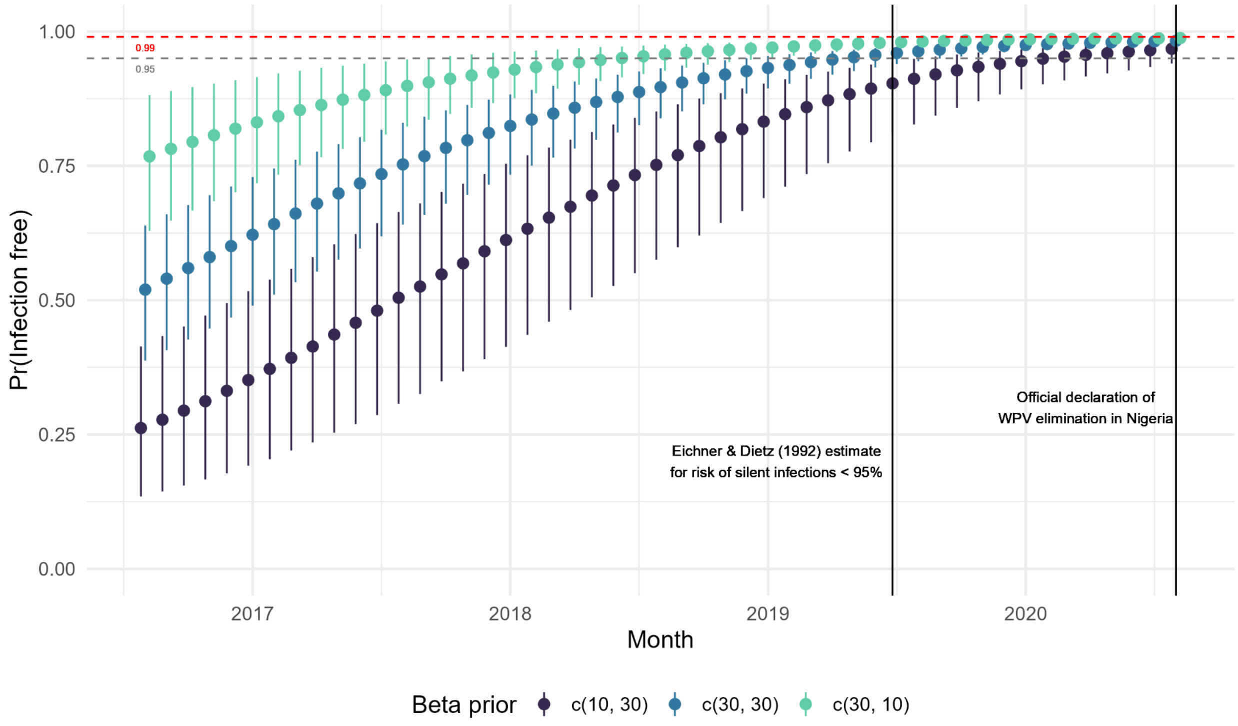


***Figure S7:*** *Estimates trajectories of the probability of freedom from infection, according to two alternative assumed distributions for the prior probability at the first time point without detection (assumed distribution for the primary analysis - a Beta distribution with* $\alpha = 30$ *and* $\beta = 30$ *- shown blue). Regardless of which prior was assumed, the estimated FFI probability had exceeded 95% before the official declaration of elimination in August 2020.*

#### Time-varying versus static sensitivity (2016-2020)

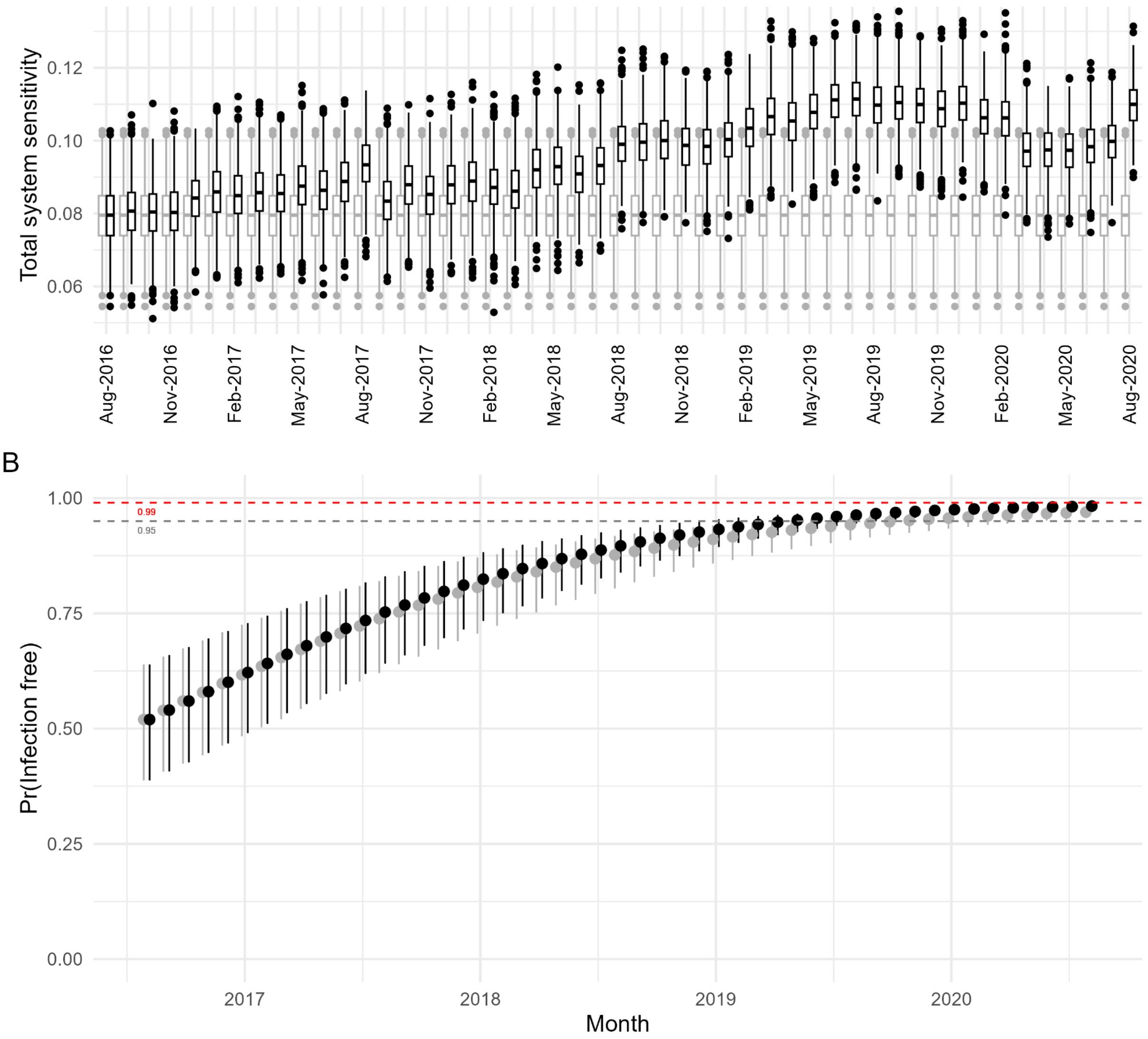


***Figure S8:*** *Comparison of inferred probability of freedom from infection, with and without taking into account varying sensitivity of surveillance over time. For the period 2016-2020, environmental surveillance was expanded and as a result the overall sensitivity of surveillance increased. Not accounting for this increase under-estimates the accumulating evidence of freedom from infection, and results in an additional five months until the FFI probability exceeds 95%.*

9. Global Polio Eradication Initiative. Field guidance for the implementation of environmental surveillance for poliovirus. World Health Organization; 2023.
